# A third vaccine dose equalizes the levels of effectiveness and immunogenicity of heterologous or homologous COVID-19 vaccine regimens

**DOI:** 10.1101/2023.02.13.23285853

**Authors:** Nicolas Guibert, Kylian Trepat, Bruno Pozzetto, Laurence Josset, Jean-Baptiste Fassier, Omran Allatif, Kahina Saker, Karen Brengel-Pesce, Thierry Walzer, Philippe Vanhems, Sophie Trouillet-Assant, Lyon-COVID study group

## Abstract

**Backgroung:** To cope with the persistence of the Covid-19 epidemic and the decrease in antibody levels following vaccination, a third dose of vaccine has been recommended in the general population. However, several vaccine regimens had been used initially, and the heterologous ChadOx1-S/BNT162b2 regimen had shown better efficacy and immunogenicity than the homologous BNT162b2/BNT162b2 regimen.

**Aim:** We wanted to determine if this benefit was retained after the third dose.

**Methods:** We combined an observational study of SARS-COV-2 infections among vaccinated healthcare workers at the University-Hospital of Lyon, France, with an analysis of immunological parameters before and after the third mRNA vaccine dose.

**Results:** Following the second vaccine dose, heterologous vaccination regimens were more protective against infection than homologous regimens, but this was no longer the case after the third dose. RBD-specific IgG levels and serum neutralization capacity against different SARS-CoV-2 variants were higher after the third dose than after the second dose in the homologous regimen group, but not in the heterologous group.

**Conclusion:** The advantage conferred by heterologous vaccination is lost after the third dose both in terms of protection and immunogenicity. Immunological measurements suggest that heterologous vaccination induces maximal immunity after the second dose, whereas the third dose is required to reach the same level in individuals with a homologous regimen.

## Introduction

In response to the coronavirus disease pandemic, several vaccines were rapidly designed and administered to the population, inducing a protective immunity composed of both neutralizing antibodies and virus-specific T lymphocytes. As multiple vaccines were available, different heterologous combinations of prime/boost doses have been used in patients. This mixing of vaccines was motivated first by the necessary adaptation to limited vaccine supply, but also by the rare observation of vaccine-induced severe adverse reactions with some vaccines. Most studies have reported similar or higher immunogenicity following heterologous primary vaccination involving the Vaxzevria (ChAdOx1nCoV-19, AstraZeneca, Cambridge, United Kingdom) and mRNA vaccines Comirnaty (BNT162b2, BioNTechPfizer, Mainz, Germany/New York, United States) and Spikevax (mRNA-1273, Moderna, Cambridge, United States) compared to homologous vaccination [1–4]. For example, binding and neutralizing antibody titers were similar or greater in the heterologous boosted group compared to the homologous group [5,6]. In addition, we and others reported that the enhanced immunogenicity of the heterologous vaccination regimen was associated with a better protection against SARS-CoV-2 infection[2,7]. However, numerous studies have shown that the level of antibodies, and in particular those neutralizing the virus, gradually decreased following vaccination. This phenomenon, combined with the emergence of viral variants having acquired mutations in the spike viral protein making them less sensitive to vaccine antibodies, led health authorities to recommend the injection of a third booster dose. This booster dose was particularly important in immunosuppressed patients for whom vaccine efficacy was lower. Heterologous vaccine schedules of ChAdOx1-S priming and mRNA booster doses as both second and third doses were not associated with increased risk of serious adverse events compared with homologous mRNA vaccine schedules [8]. Recent reports demonstrated that most COVID-19 vaccines delivered as a third dose booster significantly enhanced both humoral and cellular anti-SARS-CoV-2 immunity [9]. Observational studies also suggest that a third dose significantly improves protection from symptomatic infection compared to two doses. A recent meta-analysis reported that heterologous and homologous three-dose regimens work comparably well in preventing covid-19 infections, even against different variants [10]. Nevertheless, a recent report has documented some differences in immunogenicity and protection according to vaccine schedule before third dose [11], suggesting that initial vaccination regimens could imprint spike-specific immunity in the long term, regardless of the booster dose.

To address this question, we compared spike-specific immunity and protection against infection conferred by second and third dose of mRNA vaccine in healthcare workers (HCWs) primed with either adenovirus-based ChAdOx1-S or COVID-19 mRNA vaccine.

## Methods

### Prevalence of SARS-CoV-2 VOC

SARS-CoV-2 testing of HCWs was performed using routine diagnostic procedures in the Virology laboratory of the Hospices Civils de Lyon, and included: TMA (Aptima™ SARS-CoV-2 Assay (Hologic), LAMP (SARS-CoV-2 ID NOW™ (Abbott), and RT-qPCR with different kits (Cobas® 6800 SARS-CoV-2 assay (Roche), Panther Fusion SARS-CoV-2 assay (Hologic). To determine the prevalence of SARS-CoV-2 VOC in HCW, available positive samples with Ct<28 were sequenced using COVIDSeq (Illumina) as previously described [12]. Libraries were sequenced to 1 M paired-end reads (2×100 bp) and data were analyzed using the in-house seqmet bioinformatic pipeline (available at https://github.com/genepii/seqmet). Clades and lineages were determined on samples with genome coverage >90% using Nextclade and PangoLEARN, respectively.

### Ethical statement and cohort description

#### Population of HCW from the hospital database included in the epidemiological investigation

We extracted data from the occupational medicine database of the University Hospital of Lyon (Hospices Civils de Lyon), France. A total of 13489 HCWs working at Hospices civils de Lyon throughout the study period (12/15/21 _ 03/21/22) were included. Only subjects who (i) had never contracted COVID-19, (ii) were primed with ChAdOx1-S-nCoV-19 or an mRNA vaccine and (iii) received the second or third dose of an RNA vaccine were included in the epidemiological analysis (Supplementary Figure 1). Breakthrough infections, documented by positive RT-PCR or antigenic tests and that occurred after the 15^th^ of December 2021 and at least 7 days after vaccine injection were took into account to evaluate infection risk in different groups of subjects. Due to the mandatory SARS-CoV-2 vaccination for HCWs in France, we have no missing data regarding this variable. Moreover, the declaration of SARS-CoV-2 infection is compulsory for all staff to obtain daily allowances without loss of salary during the imposed quarantine.

The use and analysis of data from the occupational health medical file were authorized after a regulatory declaration to the National Commission for Information Technology and Civil Liberties according to the reference methodology (declaration MR004 number 20-121 of April 30^th^ 2020).

#### Population included in the immune response investigations

Eighty-eight naive HCWs for COVID-19 and vaccinated with BNT and/or ChAd and/or mRNA-1273 vaccines were included in a prospective longitudinal cohort study conducted at the Hospices Civils de Lyon. Blood sampling was performed before vaccination, before and 4⍰weeks after the second and the third dose of vaccine. The absence of previous SARS-CoV-2 infection was confirmed using the Abbott SARS-CoV-2 anti-N Ab total assay in all samples (Abbott Diagnostics, Abbott Park, Illinois, United States). Demographic characteristics and delays between doses are depicted in **Supplementary Table 1**.

### Measurement of IgG titers

Serum specimens were immediately stored at –80⍰°C after blood sampling. RBD-specific IgG antibodies were measured using bioMérieux Vidas SARS-CoV-2 IgG diagnosis kits, according to the manufacturers’ recommendations. For standardization of these assays to the first World Health Organization international standard, the concentrations were transformed into binding antibody units per ml (BAU⍰ml^−1^) using the conversion factors provided by the manufacturers.

### Live-virus neutralization experiments

A plaque reduction neutralization test (PRNT) was used for the detection and titration of neutralizing antibodies as previously described [13]. Neutralization was recorded if more than 50% of the cells present in the well were preserved. The neutralizing titer was expressed as the inverse of the higher serum dilution that exhibited neutralizing activity; a threshold of 20 was used (PRNT_50_ titer⍰≥⍰20). All experiments were performed with a subset of sera specimens collected longitudinally from 15 subjects in each group. The different viral strains that were used were sequenced and deposited at GISAID (https://www.gisaid.org/) (accession numbers EPI_ISL_1707038 19A (B.38 lineage); EPI_ ISL_1904989 Delta (B.1.617.2 lineage); and EPI_ISL_7608613 Omicron (B.1.1.529 lineage)).

### Statistical analysis

The descriptive statistics generated appropriate figures and parameters according to type of variable (i.e. continuous or categorical). The comparisons between groups were done using the Chi-square test for categorical variables and non-parametric test or student T test according to the distribution for continuous variables. The cumulative probability of COVID-19 was based on the Kaplan-Meier survival analysis and survival distributions were compared using the LogRank test. A univariate and multivariate Cox proportional hazard model was performed to identify the determinants independently associated with onset of COVID-19 according to their hazard ratio (HR) and their 95% confidence interval (95% CI). All *P*-values were two-tailed. *P*⍰<⍰0.05 was considered as statistically significant. For the immunological part, we used a multiple linear regression model, with adjustment variables (age or age groups, sex, delay, vaccination scheme).

## Results

### Vaccine effectiveness in HCWs

To compare the risk of SARS-CoV-2 infection following a second or third dose of COVID-19 mRNA vaccine (the Pfizer-BioNTech mRNA vaccine, BNT162b2, or the Moderna mRNA vaccine, mRNA-1273) in subjects who received a priming dose of ChAdOx1-S or BNT162b2 vaccine, we extracted data from the occupational medicine database of the University Hospital of Lyon (Hospices Civils de Lyon, HCL), France. We focused on individuals not previously infected with SARS-CoV-2 (PCR diagnosis) before vaccination to avoid any bias linked to hybrid immunity (**Figure 1**). Infection rates were monitored in HCWs after the second or the third booster dose in each group. We extracted SARS-CoV-2 infection incident events documented by positive antigenic or RT–PCR tests that occurred between December 15^th^ 2021 and 21^st^ March 2022. This period corresponds to the Omicron (lineage BA.1, Clade 21K) wave, succeeding to Delta (lineage B.1.1.529, Clades 21I and 21J), and preceding the Omicron sub-lineage BA.2 (Clade 21L) appearance and increase (**Figure 2A-B**). The proportion of SARS-CoV-2 variants of concern (VOC) determined by whole genome sequences in samples collected in HCWs working at Hospices Civils de Lyon during this period was: 70/834 (8.4%) 21J (Delta), 708/834 (84.9%) 21K (Omicron BA.1), and 56/834 (6.7%) 21L (Omicron BA.2). **Figure 2C-D** shows the cumulative incidence of breakthrough infections in each group. Following the second vaccine dose, heterologous vaccination regimens were more protective against infection compared to the homologous regimen group (**Figure 2C**). Indeed, after adjustment on age, sex and delay between last vaccination and the start of the study, individuals vaccinated with mRNA vaccines were twice as likely to be infected than those vaccinated with ChAdOx1-S followed by mRNA vaccine (adjusted hazard ratio of 1.88 [1.18-3.00], p-value =0.008, **Figure 2E**). After the third dose, the number of infections was lower than after the second dose in both vaccination groups, showing the benefit of the boost. Moreover, in the homologous group, the third dose achieved at least the same level of protection as in the heterologous group as demonstrated by the inversion of the infection incidence curves (adjusted hazard ratio of 0.86 [0.72 – 1.02], p-value =0.082, **Figure 2F**). We also conducted an analysis of the infection risk according to the parameters of vaccination, sex or age class. This analysis confirmed the importance of the vaccination schedule after the second dose but not after the third (**Figure 2G-H**). It also shows that in this cohort of vaccinated HCWs, middle-aged females had a higher risk of infection.

**Figure 1:**
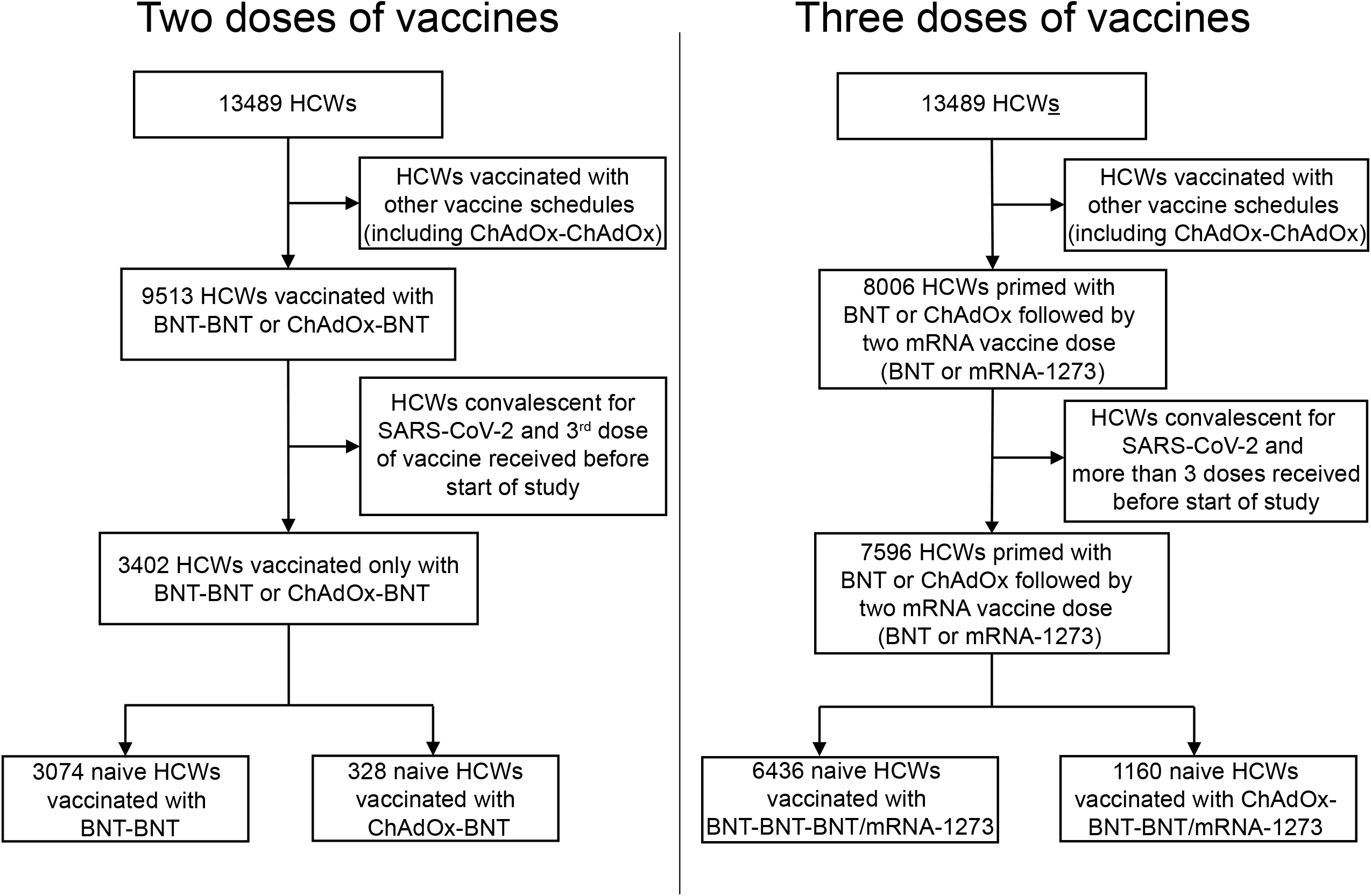
Flow chart of the epidemiological study. The flow chart indicates how individuals with two (left) or three vaccination doses (right) were selected from the total HCW population according to the indicated criteria of exclusion.

**Figure 2:**
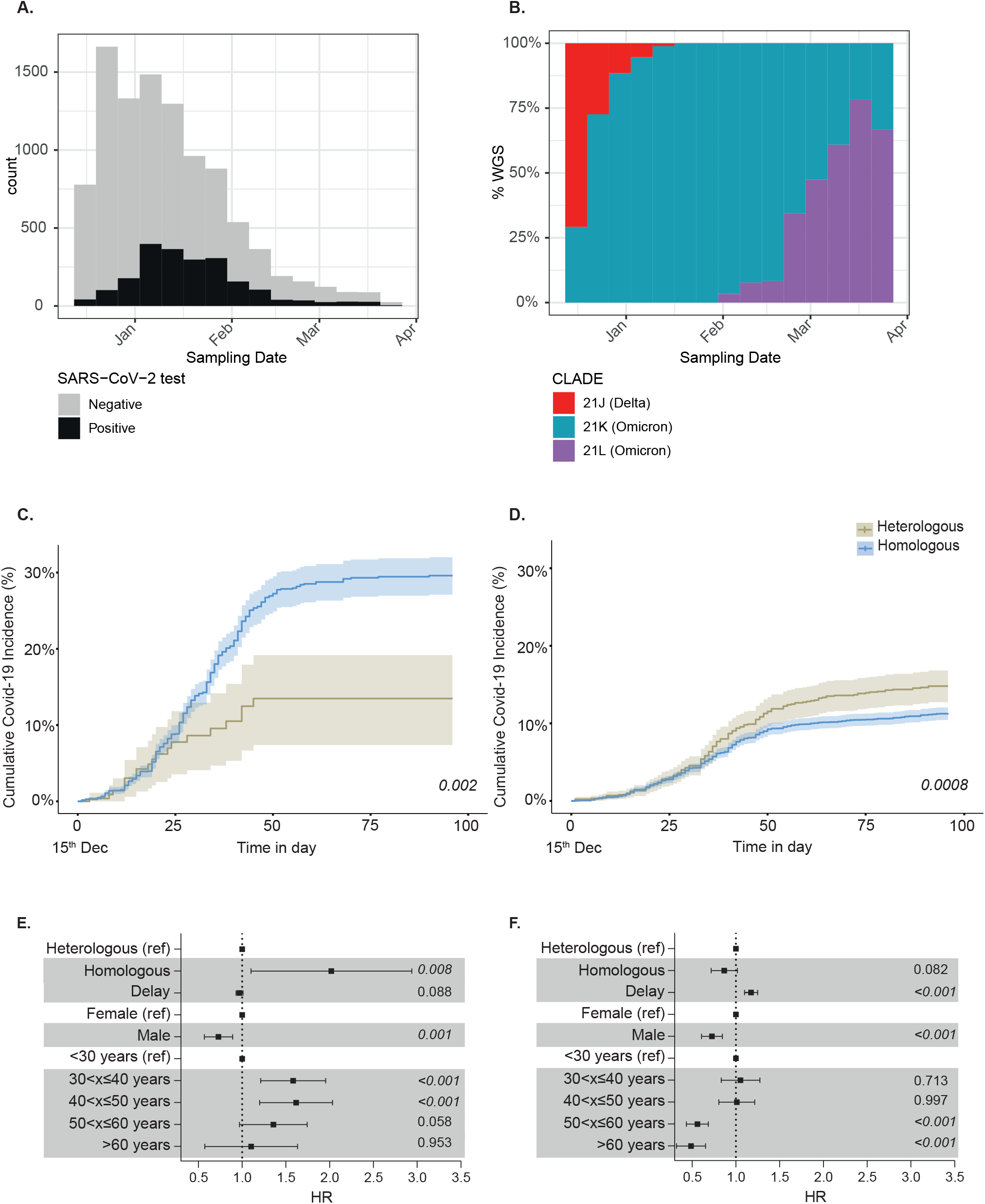
The superior efficacy of heterologous vaccination is lost after the third vaccine dose (A) Weekly SARS-CoV-2 testing (n= 2,113 positive samples / 7,863 negative samples) and (B) Variant of concern (VOC) circulation (B) among HCW (n=834 samples with whole genome sequence (WGS) between Dec 15^th^, 2021 and March 21^st^, 2022 at Lyon University Hospitals. (C-D) Graphs show the cumulative probability of COVID-19 infection within 100 days after a 2^nd^ (C) or a 3^rd^ (D) mRNA vaccine dose in health care workers at Lyon university hospital primed with BNT162b2 (blue line, homologous vaccination, 3074 and 6436 HCWs respectively) or ChAdOx1-S vaccine (brown line, heterologous vaccination, 328 and 1160 HCWs respectively). P-value was calculated using Logrank test. (E-F) Forest Plots depict multivariate Cox proportional hazard ratio (IC95%) for COVID-19 infection after a 2^nd^ (E) or a 3^rd^ (F) mRNA vaccine dose in health care workers. Hazard Ratio (HRs) were calculated with the HR of 1.0 of the reference stratum, references were indicated for all comparisons. All SARS-CoV-2 infection events documented by positive SARS-CoV-2 RT-PCR or antigenic test were recorded by the service of occupational medicine, Hospices Civils de Lyon.

### Analysis of immune response after heterologous and homologous vaccination

We then sought to compare the immunogenicity of the third dose in heterologous vs homologous vaccination groups. For this, we took advantage of the Covid-Ser cohort that we previously described and which includes a subset of voluntary healthcare workers in whom anti-SARS-CoV-2 immunity is measured longitudinally over time (**Figure 3**)[2]. We formed groups who had received a homologous or heterologous vaccination regimen and who had received a third dose of mRNA vaccine (**Table 1**).

**Figure 3:**
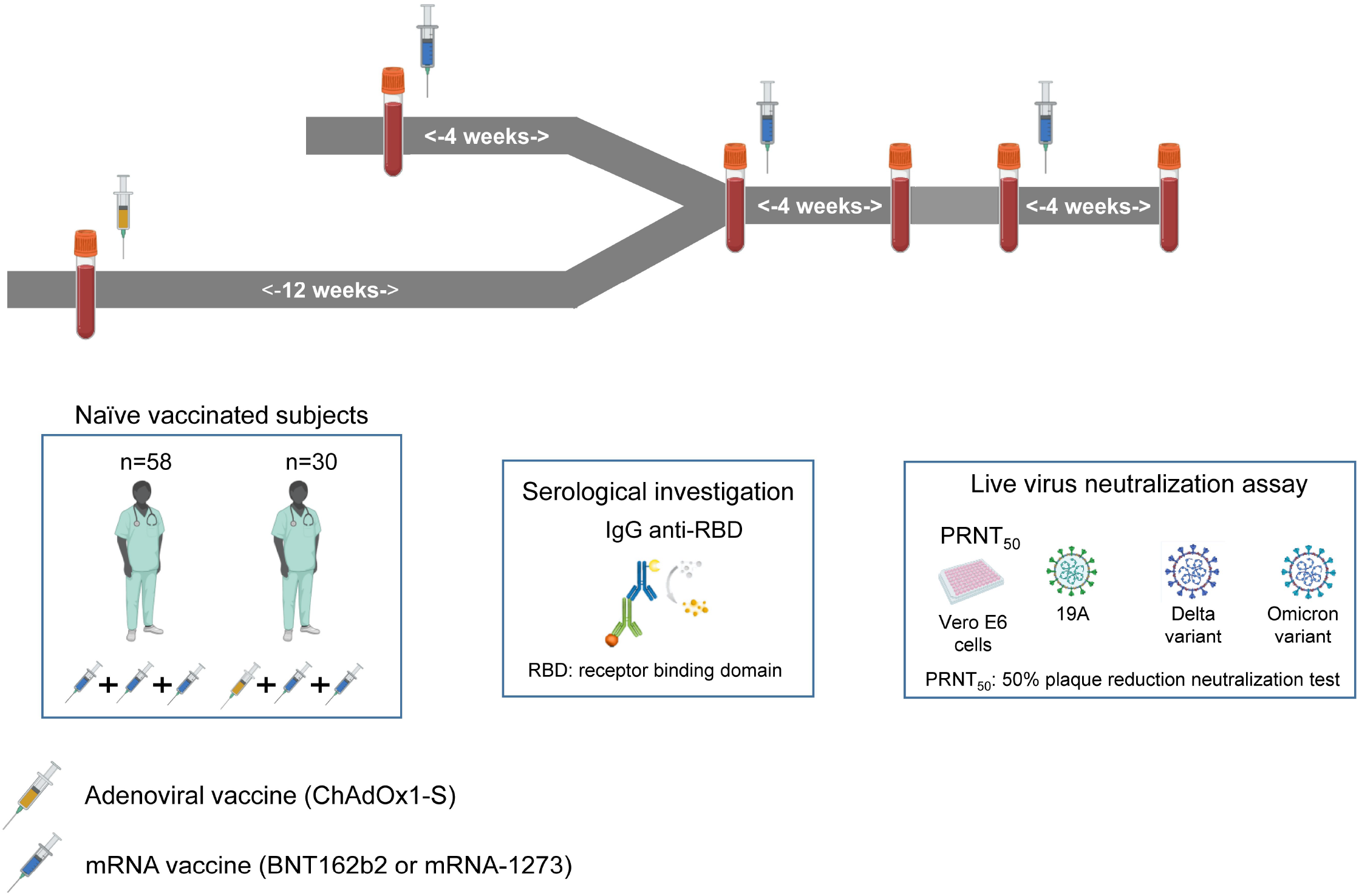
Design of the “Covid-Ser” study

**Table 1:**
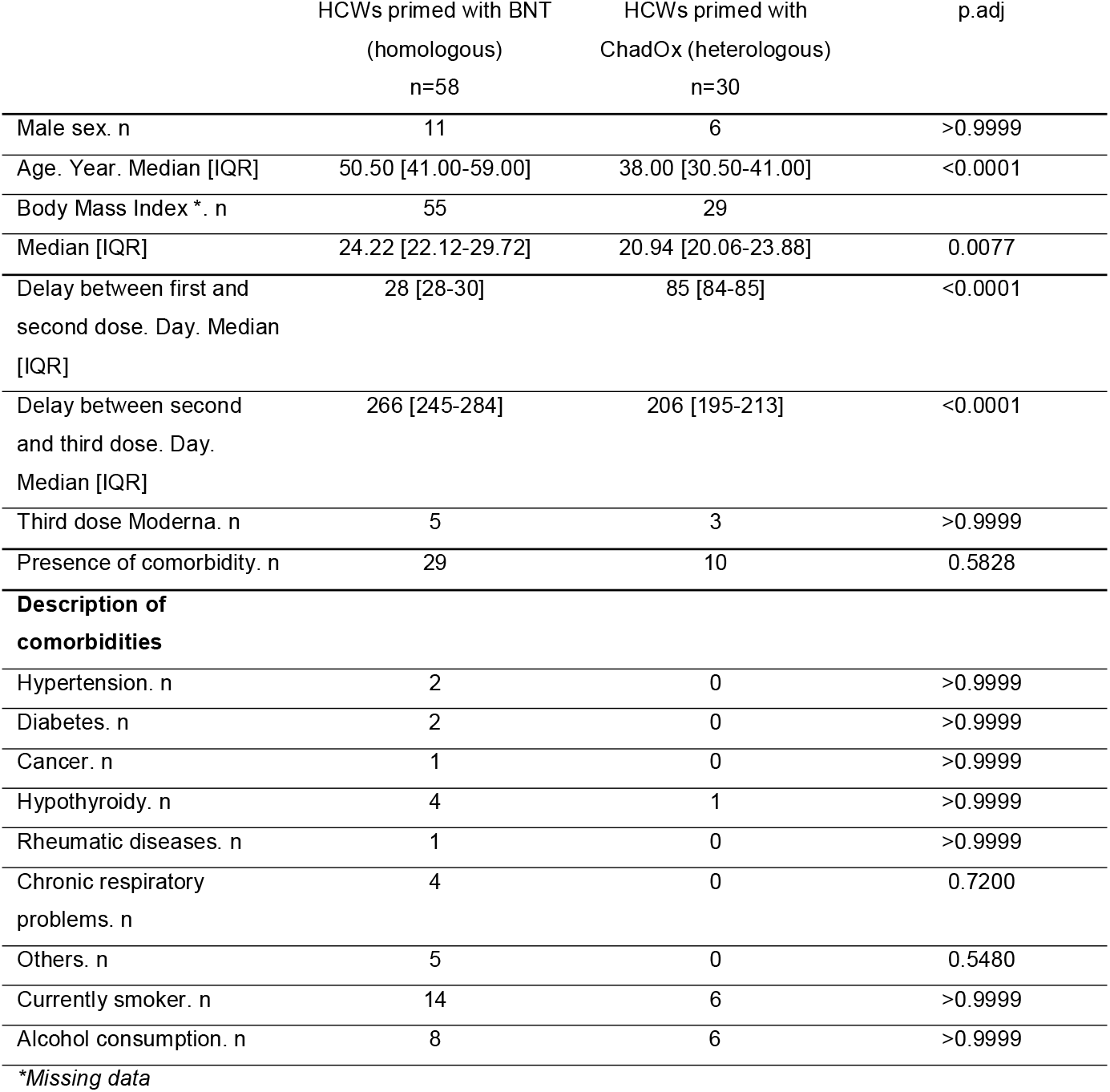
Clinical characteristics of patients in the Covid-Ser study (immunological analysis) For alcohol consumption, this was defined as consumption at least once a week. Wilcoxon-Mann–Whitney two-sided tests were used for quantitative variables and Chi-square or Fisher were used for qualitative variables when appropriate. Adjusted P values were calculated using the Benjamini–Hochberg method.

|  | HCWs primed with BNT<br>(homologous)<br>n=58 | HCWs primed with<br>ChadOx (heterologous)<br>n=30 | p.adj |
| --- | --- | --- | --- |
| Male sex. n | 11 | 6 | >0.9999 |
| Age. Year. Median [IQR] | 50.50 [41.00-59.00] | 38.00 [30.50-41.00] | <0.0001 |
| Body Mass Index *. n | 55 | 29 |  |
| Median [IQR] | 24.22 [22.12-29.72] | 20.94 [20.06-23.88] | 0.0077 |
| Delay between first and<br>second dose. Day. Median<br>[IQR] | 28 [28-30] | 85 [84-85] | <0.0001 |
| Delay between second<br>and third dose. Day.<br>Median [IQR] | 266 [245-284] | 206 [195-213] | <0.0001 |
| Third dose Moderna. n | 5 | 3 | >0.9999 |
| Presence of comorbidity. n | 29 | 10 | 0.5828 |
| **Description of**<br>**comorbidities** |  |  |  |
| Hypertension. n | 2 | 0 | >0.9999 |
| Diabetes. n | 2 | 0 | >0.9999 |
| Cancer. n | 1 | 0 | >0.9999 |
| Hypothyroidy. n | 4 | 1 | >0.9999 |
| Rheumatic diseases. n | 1 | 0 | >0.9999 |
| Chronic respiratory<br>problems. n | 4 | 0 | 0.7200 |
| Others. n | 5 | 0 | 0.5480 |
| Currently smoker. n | 14 | 6 | >0.9999 |
| Alcohol consumption. n | 8 | 6 | >0.9999 |

\*Missing data

Subsequently, we measured the increase in anti-RBD IgG antibody levels four weeks after the third dose in both groups. No significant difference was observed (p=0.14) (**Figure 4A**). We then evaluated the neutralization capacity of serum antibodies against SARS-CoV-2 variants 19A, Delta and Omicron four weeks after the third dose. No significant difference in neutralization was observed between the homologous and heterologous schedules (19A: median [IQR] 960 [320-1920] vs. 640 [320-1920]; Delta: 240 [120-480] vs. 320 [160-960]; Omicron: 160 [60-640] vs. 160 [80-480], respectively) (**Figure 4B**). Moreover the anti-RBD IgG level is not different in the two groups after the third dose (p=0.18) (**Figure 4C**).

**Figure 4:**
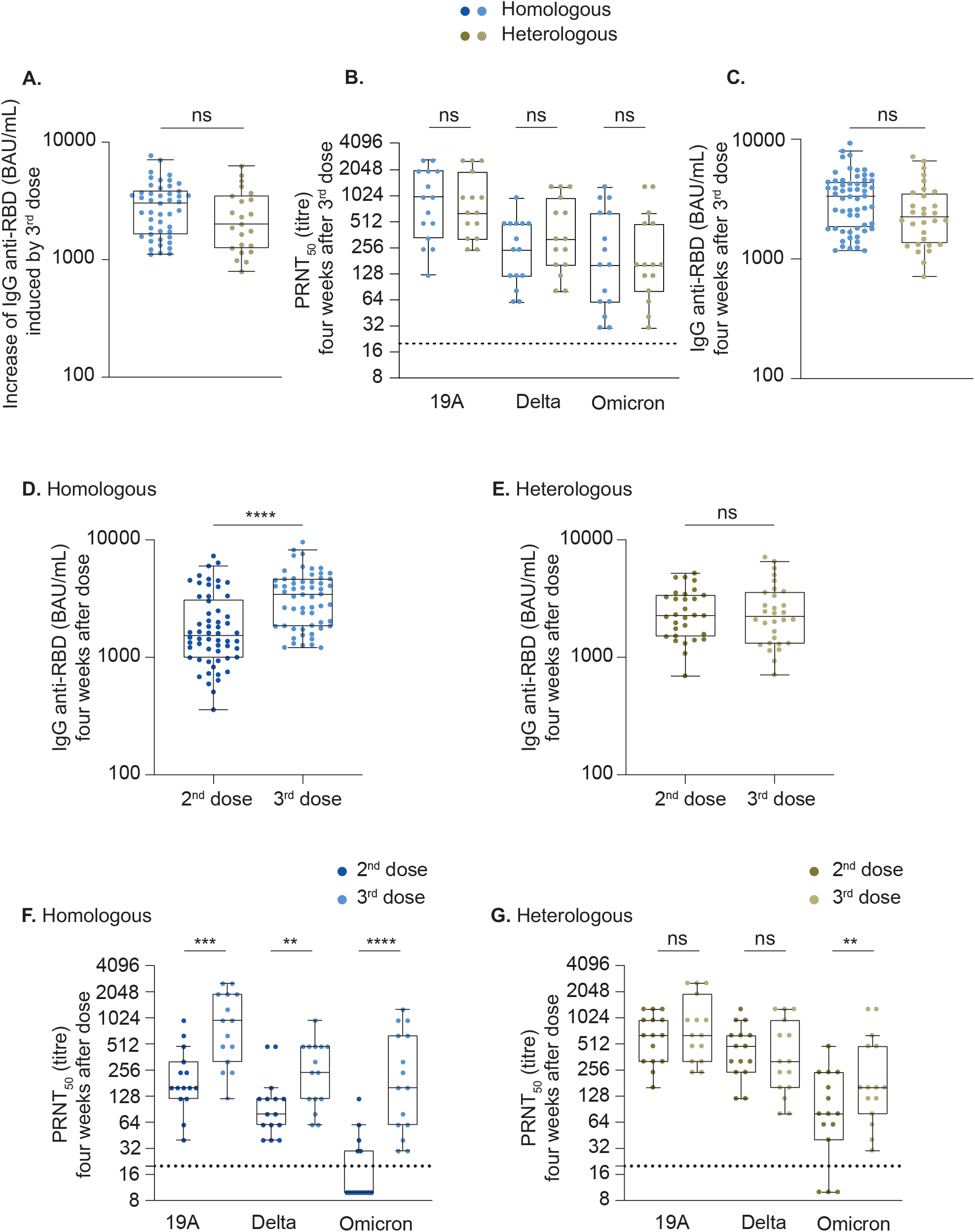
The booster dose equalizes the levels of immunogenicity of heterologous or homologous vaccine regimens Sera from naïve HCWs vaccinated primed with the BNT162b2 (n=58) or ChadOx1-S vaccine, n=30) and who received an mRNA vaccine for 2^nd^ and 3^rd^ dose were selected. (A) RBD-specific IgG levels were quantified just before and four weeks post booster (third) dose. Concentrations are expressed in binding antibody units per ml (BAU/ml), and data show the difference in RBD-specific IgG levels between the two time-points (missing value for pre-vaccination time point for 11 and 4 patients vaccinated with homologous and heterologous regimen respectively). (B) A subset of 15 sera from both groups four weeks after the third dose were assayed in duplicate for their capacity to neutralize infection of Vero E6 cells by different SARS-CoV-2 strains, as indicated. (C) Comparison of serum RBD-specific IgG levels 4 weeks after the 3^rd^ dose between homologous and heterologous vaccine recipients (D-E) Serum RBD-specific IgG levels were measured 4 weeks after the 2^nd^ or after the 3^rd^ dose in the homologous (D) or heterologous (E) vaccine recipients. (F-G) Comparison of the neutralizing capacity of total antibodies collected 4 weeks after the 2^nd^ or after the 3^rd^ dose in the homologous (F) or in the heterologous (G) vaccine group (n=15 for each group). The dashed lines represent the limit of detection (20 in PRNT_50_). In all graphs, data are represented as box-and-whiskers plots using the Tukey method which show median (horizontal line inside the box), interquartile (25%-75% - upper and lower horizontal lines of the box), and each dot corresponds to one subject. Statistics were calculated using a multiple linear regression model with adjustment variables (age and sex, and when appropriate delay between second and third dose).

Data in **Figure 2** show that the heterologous ChAdOx1-S/COVID-19 mRNA vaccine combination confers better protection against SARS-COV-2 infection than the homologous COVID-19 mRNA vaccines combination, but that the third dose equalizes the efficacy of both vaccine regimens. To understand why the advantage of heterologous vaccination is no longer observed after the third dose, we compared humoral immunity four weeks after the second and after the third dose in each group. In subjects vaccinated according to the homologous regimen, the anti-RBD IgG level measured 4 weeks post second dose was significantly lower (p<0.0001) than that measured post third dose (1490 [969-2994] BAU/mL, 3336 [1795-4491] BAU/mL respectively) (**Figure 4D**) whereas in subjects vaccinated according to a heterologous scheme, no difference was observed (2277 [1520-3400] BAU/mL vs 2242 [1321-3602] BAU/mL respectively) (**Figure 4E**). In addition, for the homologous vaccine group, the antibody neutralizing capacity 4 weeks post third dose was at least 3 times higher than that observed 4 weeks post second dose (at least p<0.01). In contrast, in heterologous vaccinated individuals, no benefit in neutralizing capacity against 19A and Delta at 4 weeks post third dose was observed. Only the neutralizing capacity of the total antibodies 4 weeks post third dose against Omicron increased 2-fold compared to that 4 weeks post second dose (p<0.01) (**Figure 4F-G**). Of note, there is no difference of anti-SARS-CoV-2 IgG level 4 weeks post third dose between HCWs boosted with BNT162b2 (n=80) or mRNA-1273 (n=8) vaccine (p=0.30).

## Discussion

We previously showed that the heterologous ChAdOx1-S/BNT162b2 combination confers better protection against SARS-CoV-2 infection than the homologous BNT162b2/BNT162b2 combination in a real-world observational study conducted in HCWs [2]. Both combinations induced strong anti-spike antibody responses but serum specimens from heterologous vaccinated individuals displayed a stronger neutralizing activity, regardless of the SARS-CoV-2 variant [14]. Here, we asked whether the advantage conferred by the heterologous regimen is conserved after a booster dose of mRNA-based COVID-19 vaccine. Our results show that (i) the third dose with an mRNA vaccine equalizes the levels of effectiveness of heterologous or homologous COVID-19 vaccine regimens and (ii) that serum neutralization capacity against different SARS-CoV-2 variants is comparable four weeks after the boost in both groups. Indeed, the third vaccine dose does not increase antibody levels and neutralization capacity beyond those observed one month after the second dose in the heterologous group, which suggests that a maximal immunity level one month post vaccination is already reached after the second dose in this group and that it cannot be boosted further, at least with an mRNA-based vaccine. While we cannot exclude that a shorter delay between the second and third doses would have a different impact in heterologous vs homologous groups, our results are in line with those of Accorsi et al. who showed that a single booster dose of an mRNA Covid-19 vaccine in individuals who received primary vaccination with a single-dose of adenovirus-based vaccine Ad26.COV2.S, provided protection close to that of the three-dose mRNA vaccine regimen [15]. In addition, Behrens et al. reported that inferior SARS-CoV-2 specific immune responses following homologous ChAdOx1-S / ChAdOx1-S vaccination compared to ChAdOx1-S /BNT162b2 can be compensated by heterologous BNT vaccination as third dose [16]. In our study, only the neutralization of the Omicron variant was slightly better one month after the third dose in the heterologous regimen compared to the second dose. The significance of this result is not clear since the neutralization of the other variants was not changed.

More largely, our results address the question of the number of vaccine boosts that are needed to reach the maximal immunity level against SARS-CoV-2, including its numerous variants. We confirm that, using the heterologous combination performed in our study, a single boost is enough to reach this plateau. By contrast, the homologous scheme using an mRNA vaccine needs two boosts for reaching the same level of protection. Of course, this maximum immunity is temporary and decreases with time, which makes the third dose necessary in the heterologous group as well. The recent study of Regev-Yochay et al. that evaluated the benefit of a third boost in an homologous mRNA immunization scheme suggests that a maximum immunity is reached after the third dose with homologous mRNA vaccination. The fourth mRNA vaccine dose seems to be able to restore the level of immunity, but does not quantitatively and qualitatively improve the humoral immunity conferred by the first 3 doses [17].

Surprisingly, our data highlighted a higher risk of infection in women between 30 and 50 years compared to older HCW. This group of individuals mainly corresponds to active nursing staff with higher exposure to pathogens. Indeed, in the Lyon University hospital, the pattern of contacts (frequency and duration) between nurses and patients and within nurses is superior than for other professionals [18], and this pattern could result in a higher exposure to pathogens, as previously reported in a study of nosocomial influenza spreading at the hospital [19]. Obviously, this higher exposure of middle-aged women is specific to the HCW community and may not apply to the general population.

Some limitations of the present study should be acknowledged. First, these results were obtained from observational data and not from a randomized clinical trial. As a result there were some inherent differences between the compared groups. For example subjects vaccinated with a heterologous schedule were on average younger than those vaccinated with a homologous schedule. This difference is explained by the recommendations for vaccination according to which individuals under 55 years of age who received a first dose of ChAdOx1-S should receive a boost of COVID-19 mRNA vaccine. Yet, our statistical analysis did not show a significant impact of age in infection risk or vaccine-induced immune parameters. Second, the HCWs might slightly differ from the general population since they exhibit repeated exposures to SARS-CoV-2, together with close monitoring regarding vaccine coverage and COVID-19 incidence. Third, in our study, the advantage of the heterologous regimen observed after the second dose may be impacted by the delay between the first and second dose which was only of 4 weeks between the first two doses in the homologous scheme. A study by Payne et al. reported that an extended delay of 10 weeks between the first two doses of BNT162b2 allowed the development of better humoral immunity [20]. Fourth, the present study was limited to HCWs having no history of SARS-CoV-2 infection prior to vaccination. The benefit of booster dose of mRNA vaccine in patients primed with ChAdOx1-S or mRNA vaccine would need further investigations in people previously infected by different variants. Finally, even if HCWs were heavily tested during the monitoring period of our study, asymptomatic infections may have remained unnoticed.

Taken together, our data provide evidence to understand the number of vaccine boosts that are needed to reach the maximal immunity level against SARS-CoV-2 in heterologous or homologous vaccine scheme. More studies will be needed to determine if another vaccine type should be given to boost even further SARS-CoV-2 immunity.

## Data Availability

All data produced in the present study are available upon reasonable request to the authors

## Ethical statements_

For the Covid-Ser study, clinical data were recorded by a trained clinical research associate using Clinsight software (v.Csonline 7.5.720.1). Written informed consent was obtained from all participants. Ethics approval was obtained from the national review board for biomedical research in April⍰2020 (Comité de Protection des Personnes Sud Méditerranée I, Marseille, France; ID RCB 2020-A00932-37), and the study was registered at ClinicalTrials.gov (NCT04341142).

